# The First Report of a Homozygous *TUBG1* Splice Variant: Expanding the Genetic Spectrum of Tubulinopathies and Confirming the Dominant-Negative Effect

**DOI:** 10.1101/2025.03.11.25323037

**Authors:** Badreddine Elmakhzen, Yongjun Song, oussama Kettani, Yasser Ali El Asri, Zineb Rchiad, Laila Bouguenouch, Omar Askander

## Abstract

Tubulinopathies, or tubulin-related cortical dysgenesis, are a group of overlapping brain disorders caused by pathogenic mutations in tubulin-encoding genes. These disorders include microcephaly, polymicrogyria, and lissencephaly/pachygyria, which are commonly associated with severe seizures and intellectual incapacity. *TUBG1*, which encodes tubulin-gamma-1, is critical for microtubule nucleation at the microtubule organizing center (MTOC), which is required for neuronal development. Previously, only autosomal dominant missense variants of *TUBG1* were reported, which had been predicted to act through a dominant-negative mechanism. We provide the first case of a homozygous splice variation in *TUBG1* in two siblings who exhibit clinical symptoms compatible with tubulinopathy. This study increases the mutational spectrum of *TUBG1* and proposes a unique recessive inheritance mechanism. Further research is needed to confirm the pathogenicity of this variation and understand the genotype-phenotype relationship.

## Introduction

Tubulinopathies, also known as tubulin-related cortical dysgenesis, are characterized by a broad spectrum of overlapping brain abnormalities and other clinical characteristics that arise from pathogenic variations in genes that encode distinct tubulin isotypes ^1,2^. In OMIM, the diseases are classified under the phenotype series PS614039, which includes other genetic diseases related to the microtubule. These conditions include microcephaly, polymicrogyria, and lissencephaly/pachygyria. Many have been linked to severe seizures and intellectual disability; they have historically been categorized according to the biological process (migration, cortical organization, and proliferation) most likely to be impacted ^3^.

*TUBG1*, which encodes tubulin-gamma-1, a member of the tubulin superfamily, is known to cause an autosomal dominant disorder causing ‘Cortical dysplasia, complex, with other brain malformations 4 (OMIM:615412)’ ^4^. TUBG1 is a critical component of the gamma-tubulin ring complex (γ-TuRC) which mediates microtubule nucleation at the microtubule organizing center (MTOC), essential for proper cell division and neuronal development ^5^. Multiple missense variants were reported to cause the disorder in autosomal dominant inheritance, suspected of dominant-negative effect. However, no homozygous variant has been reported to this day.

Here, we present the first reported case with a homozygous splice variant in the TUBG1 gene in two siblings presented with the typical symptoms of tubulinopathy

## Methods

### Exome sequencing

Exome sequencing was performed by 3billion Inc., Seoul, South Korea. Genomic DNA was extracted from EDTA blood specimens using a standard protocol. Exome capture was performed using xGen Exome Research Panel v2 (Integrated DNA Technologies, Coralville, Iowa, USA) and sequencing was performed using NovaSeq 6000 (Illumina, San Diego, CA, USA). Generated sequences were aligned to the Genome Reference Consortium Human Build 37 (GRCh37) and Revised Cambridge Reference Sequence (rCRS) of the mitochondrial genome.

Sequencing data analysis and variant interpretation were performed using 3billion’s proprietary system, EVIDENCE v4.2 ^6^. EVIDENCE incorporates bioinformatic pipelines for calling SNV/INDEL based on the GATK best practices and Mutect2 v4.4.0 for SNV/INDEL variant calling ^7^. Manta v1.6.0 ^8^ and 3bCNV v2.1, an internally developed tool were used for CNV variant calling. ExpansionHunter v5.0.0 ^9^ were used for repeat expansion variants, MELT v2.2.2 ^10^ for calling mobile element insertion variants, AutoMap v1.2 ^11^ for detecting regions of homozygosity (ROH). Variant Effect Predictor v104.2 ^12^ is used for variant annotation. Variants were prioritized based on the guideline recommended by the American College of Medical Genetics and Genomics (ACMG) and the Association for Molecular Pathology (AMP) ^13–15^ in the context of the patient’s phenotype, relevant family history, and previous test results provided by the ordering physician.

### Sanger sequencing

Primers for the confirmation of the variant in the family members were designed using primer3.

PCR reactions were performed in a total volume of 25 µL containing 10 ng of DNA for exons of *TUBG1* gene, 100 ng of DNA for the rest of exons, 2.5 µL of 10× enzyme buffer, 0.2 mM of each dNTP, 1.5 mM MgCl2, 0.4 µM of each primer, and 0.5 U Taq DNA polymerase (Invitrogen).

Direct sequencing was performed, with the same primer sets used for PCR, on an ABI 3500Dx Genetic Analyzer v2.3 using the BigDye Terminator v3.1 chemistry (Applied Biosystems, Foster City, CA, USA).

### cDNA sequencing

The blood of the family members was collected using PAXgene tubes. mRNA of each member was extracted using PAXgene Blood RNA Kit following the manufacturer’s instruction. The mRNA was reverse transcribed using QuantiTect Reverse Transcription Kit and following the protocol. Sanger sequencing was performed with primers (F:5’-CCTAAGAAGCTGGTGCAGACA-3’, R:5’-GTCCACCTCTCCCTGGATGA-3’).

### Protein modeling

In silico prediction of the impact of the variant

Amino acid sequences of related proteins have been retrieved from UniProt ^16^. The canonical sequence of TUBG1(TBG1_HUMAN), GCP2(GCP2_HUMAN), GCP3(GCP3_HUMAN), and the TUBG1 sequence with amino acid 232-281 removed were used. The location of the deletion was visualized by PyMOL ^17^ using the structure of the gamma-tubulin ring complex(γ-TuRC) ^18^.

## Result

### Clinical presentation

Probands are children of consanguineous parents from Morocco (Figure 1). They were referred to the department of genetics in the university hospital HASSAN2 FEZ, Morocco. They had an uncle affected with mental retardation and microcephaly. The family consented for a genetic study approved by the relevant ethics committees and institutional review boards of the Fez University Hospital Ethics Committee, with ethics committee approval given to this study under number 02/2023 in accordance with the Declaration of Helsinki. (IRB code 00012098-FWA: No. 00018699. Ethics approval number: 02/2023).

Proband I was a boy in his 10’s, who was born at term by spontaneous vaginal delivery. At birth, he had a normal birth weight and was diagnosed with microcephaly (−2 SD). During infancy, He showed delayed psychomotor development and language difficulties. Before the age of 5 years old, he experienced his first epileptic seizure, which led to a diagnosis of epilepsy. He was treated with Valproate to manage the seizures. Despite these challenges, he was described as sociable and engaged in special education to support his development. He presented with dysmorphic features, including divergent strabismus and hypertelorism (contact the corresponding author to request access to these materials). At the adolescence age, he continued to have delayed psychomotor development and was diagnosed with intellectual disability. He also presented micropenis without cryptorchidism with hypotrophic testicles in ultrasound a (left:21×12×11mm, right:14×11×17 mm). Hormonal testing objectified a low level of testosterone, which was at 0.39ng/ml, FSH was at 1.63mU/ml and LH at a level of 2mU/ml.

His brain MRI revealed ribbon heterotopia in the left side associated with lateral ventricular asymmetry predominant in the left side (Figure 2F). He required ongoing follow-up for his epileptic seizures and continued to be on Valproate therapy.

Proband II, the sister of Proband I, was a girl in her 10’s who was born at term by spontaneous vaginal delivery. Her birth was also normal, with a normal birth weight, but she too was diagnosed with microcephaly at birth. Like her brother, she exhibited hypertelorism and divergent strabismus, alongside her microcephaly (images:contact the corresponding author to request access to these materials). In her early developmental years, she showed delayed psychomotor development with significant language impairment. She faced comprehension and concentration problems, which contributed to school failure and she had a slight mental retardation in comparison to her brother who was more severely delayed. Despite these challenges, she did not experience epileptic seizures, differentiating her condition from her brother’s. She was receiving educational support due to her school failures and developmental delays.

Both siblings exhibited significant neurological and developmental challenges from birth, characterized by microcephaly and delayed psychomotor development. Proband, I had additional complications with epileptic seizures and required ongoing treatment with Valproate. In contrast, proband II did not experience seizures. The MRI findings indicated ribbon heterotopia on the left side, a crucial structural brain anomaly associated with his neurological symptoms. Their clinical features included hypertelorism and divergent strabismus, which, along with microcephaly, contributed to their developmental and educational challenges (Table 1).

**Table 1.** Phenotype presentation of the probands. Both probands had microcephaly, lissencephaly, absent speech, and dysmorphic features. Only proband I had epilepsy.

|  | Proband I | Proband II |
| --- | --- | --- |
| Sex | Male | Female |
| Age at examination (year) | 5-10 | 10-15 |
| Microcephaly | Present, -2SD | Present, -2SD |
| Strabismus | Present | Present |
| Lissencephaly | Absent | Absent |
| Corpus callosum | Normal | Normal |
| Intellectual disability | Severe | Moderate |
| Motor and cognitive impairment | Present, Absent speech | Present, Absent speech |
| Epilepsy | Present | Absent |
| Radiological findings | ribbon heterotopia in the left side, enlarged ventricles | NA |

**Figure 1.**
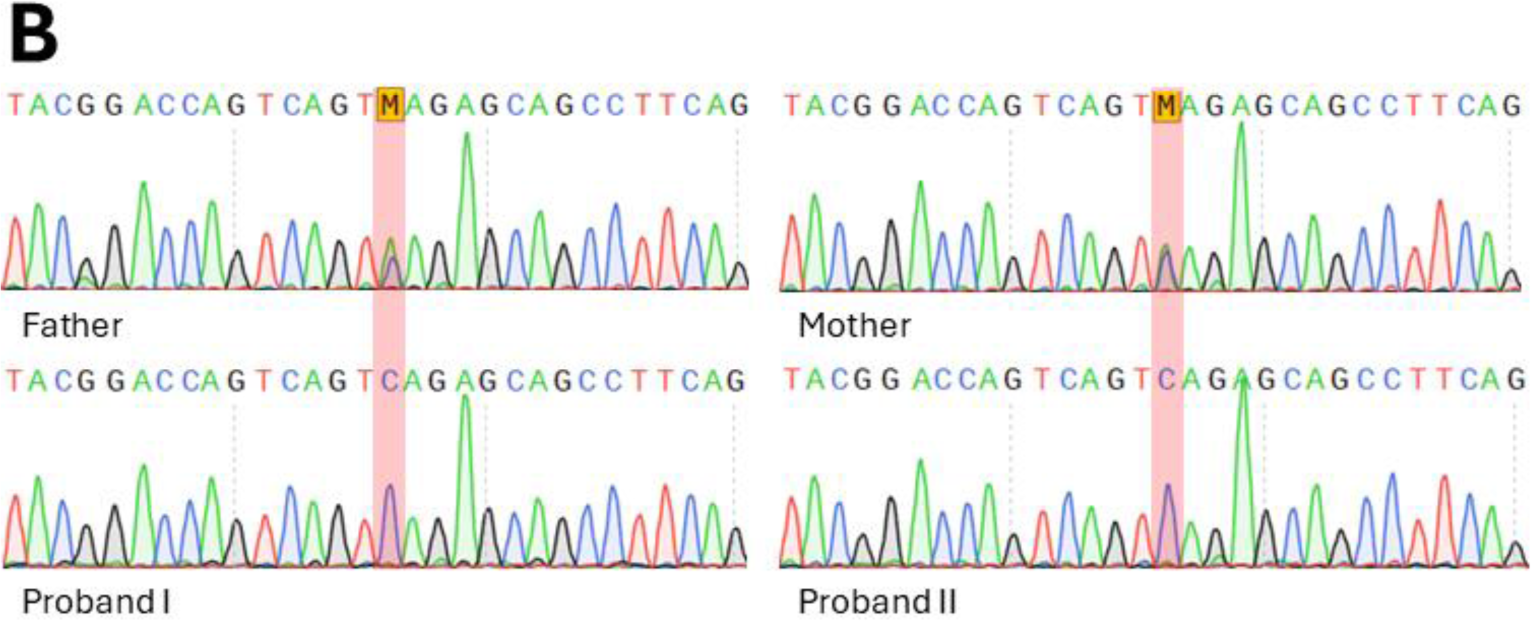
A. Pedigree of the family (upon request). Genetic tests were not performed for other family members except two probands and their parents. Arrow: Proband I, Question Mark: an uncle with similar phenotypes. B. Sanger sequencing confirmation of the four family members. Father, het / Mother, het / Sister, hom / Proband, hom

**Figure 2.**
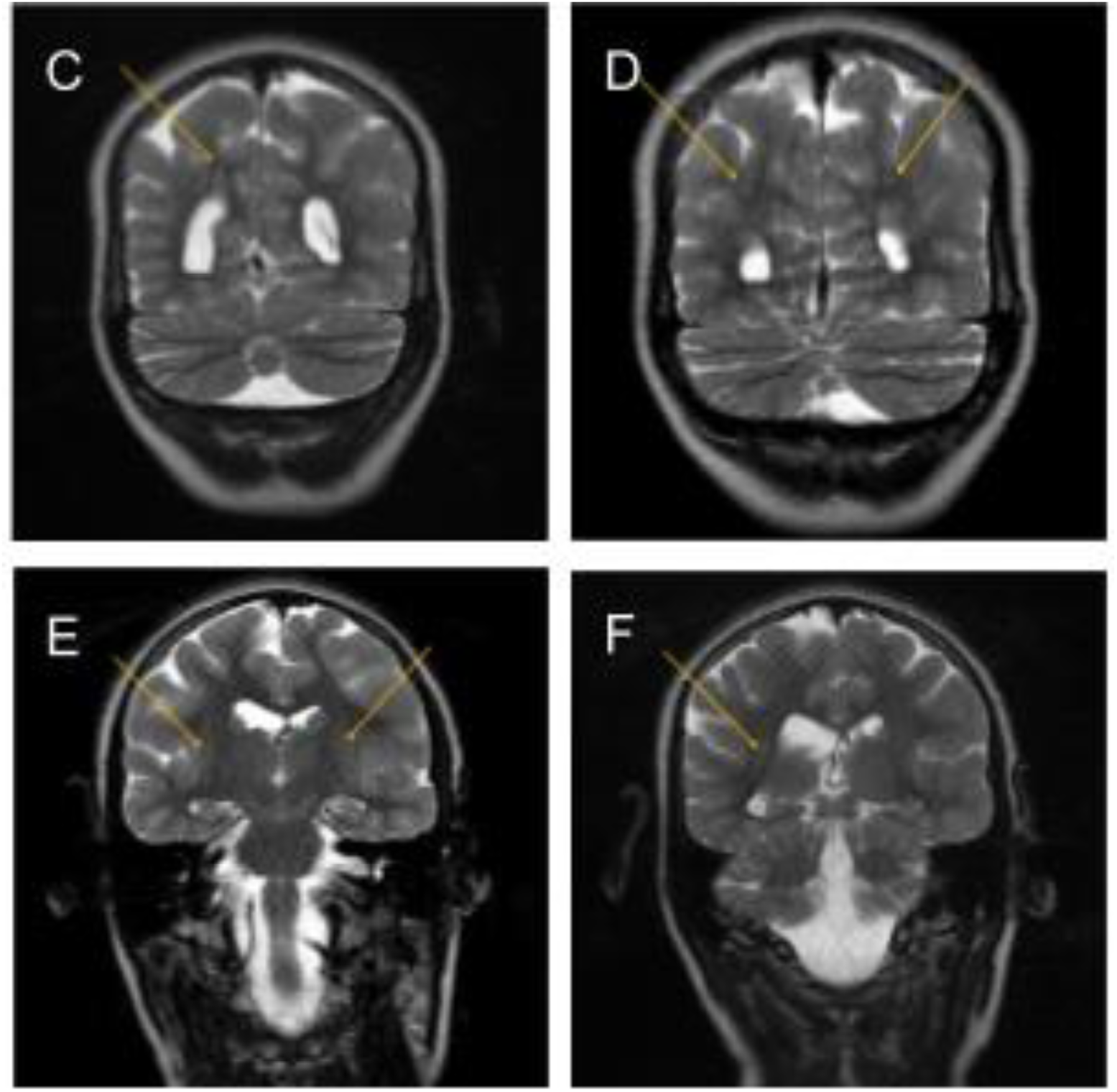
: A: Frontal images of Proband I, B: lateral images of Proband I,(upon request) Microcephaly, strabismus are present. Brain MRI images proband I : C: T2 coronal section, right periventricular nodular heterotopia, D: T2 coronal section with bilateral periventricular nodular heterotopia. E: T2 coronal section: bilateral subcortical band heterotopia. F: T2 coronal section: subcortical band heterotopia in right periventricular

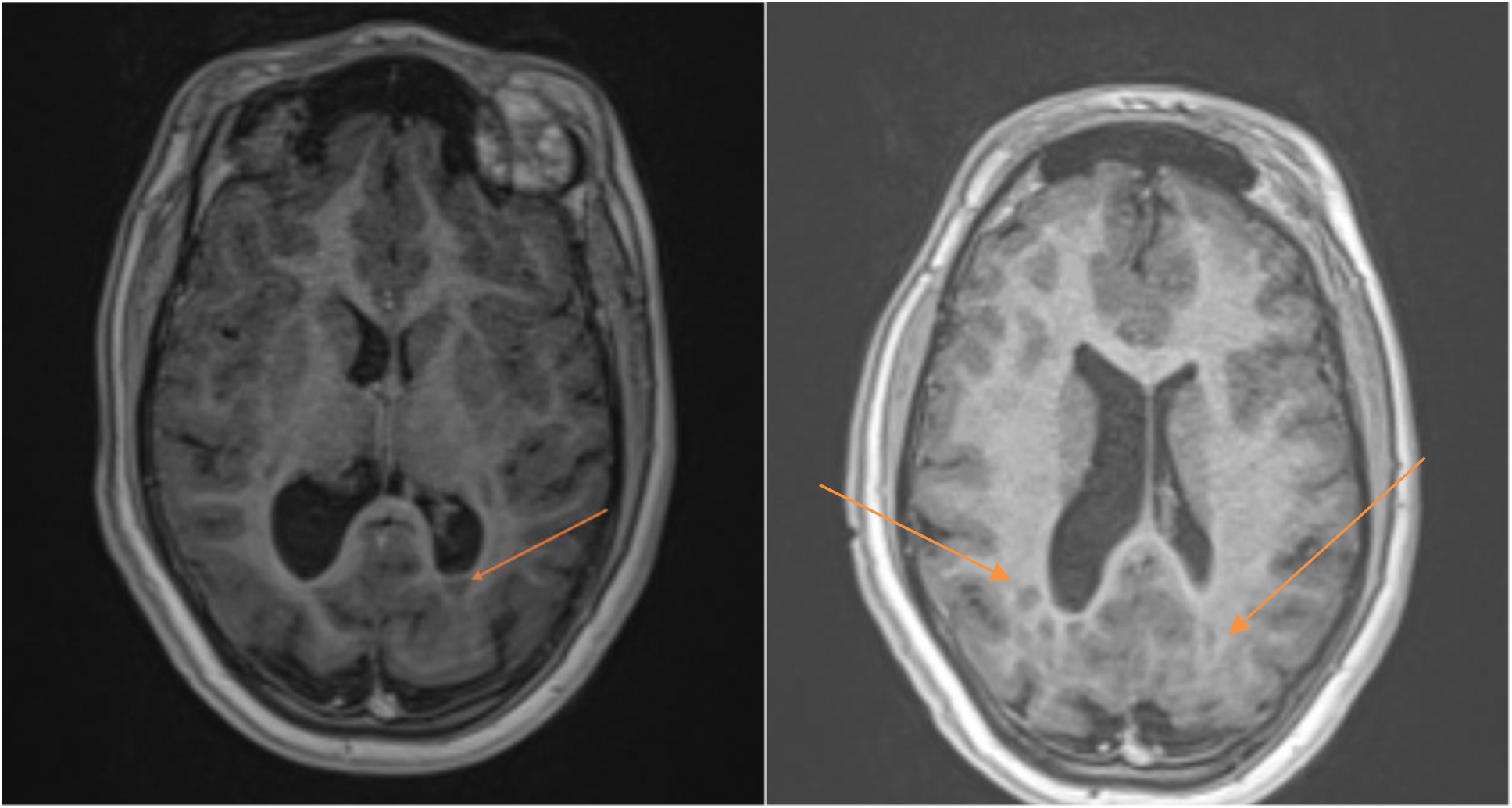
Brain MRI images: 3D T1 axial cross-sections showing nodular heterotopy of periventricular Gray matter(arrows).

### Exome sequencing of proband I and Sanger sequencing of the family

Exome sequencing was performed with the specimen of proband I. In total, 10,520,466,647 bases of sequence were generated uniquely, generating 148.57 mean depth-of-coverage within the 34,366,188 bases of the captured region, which is approximately 99.3% of the RefSeq protein coding region. 99.10% of the target region at least 20X depth-of-coverage. 72,438 single nucleotide variants (SNV) and 11,710 small insertions and deletions (indel) were identified. The result showed 2.1% of the region of homozygosity (ROH) which is compatible with the pedigree of parents being consanguineous. No clinically significant variant was discovered from disease-causing genes. Tubulinopathy-related genes (MIM: PS614039) were examined, however, variants compatible with known diseases were not discovered, besides a homozygous intron variant of uncertain significance, discovered in the TUBG1 gene. The variant, NM_001070.5:c.843+3A>C, was predicted to cause donor loss with a score of 0.92 by spliceAI resulting in exon skipping. Sanger sequencing of family members was performed. Both parents were heterozygous carriers of the variant, and the affected sibling, proband II had the same homozygous variant (Figure 1B).

### Confirmation of exon skipping

mRNA of all four family members were extracted from blood in PAXgene tubes. cDNA was generated using extracted mRNA and amplified to confirm the exon skipping. The gel electrophoresis result of PCR was compatible with the genotype of each family member. Additional Sanger sequencing was performed to identify the exact sequence of the alteration. The result showed skipping of exon 8, transcript NM_001070.5, predicted to be deletion of 50 amino acids, NP_001061.2:p.Val232_Ser281del.

### Location of the residue in 3D model

The location of the deletion was visualized using a structure of a γ-TuRC discovered by Wieczorek et al ^18^ (Figure 3). Residues removed by this deletion were located in a region where gamma-tubulin interacts with gamma-tubulin complex components (GCPs).

**Figure 3.**
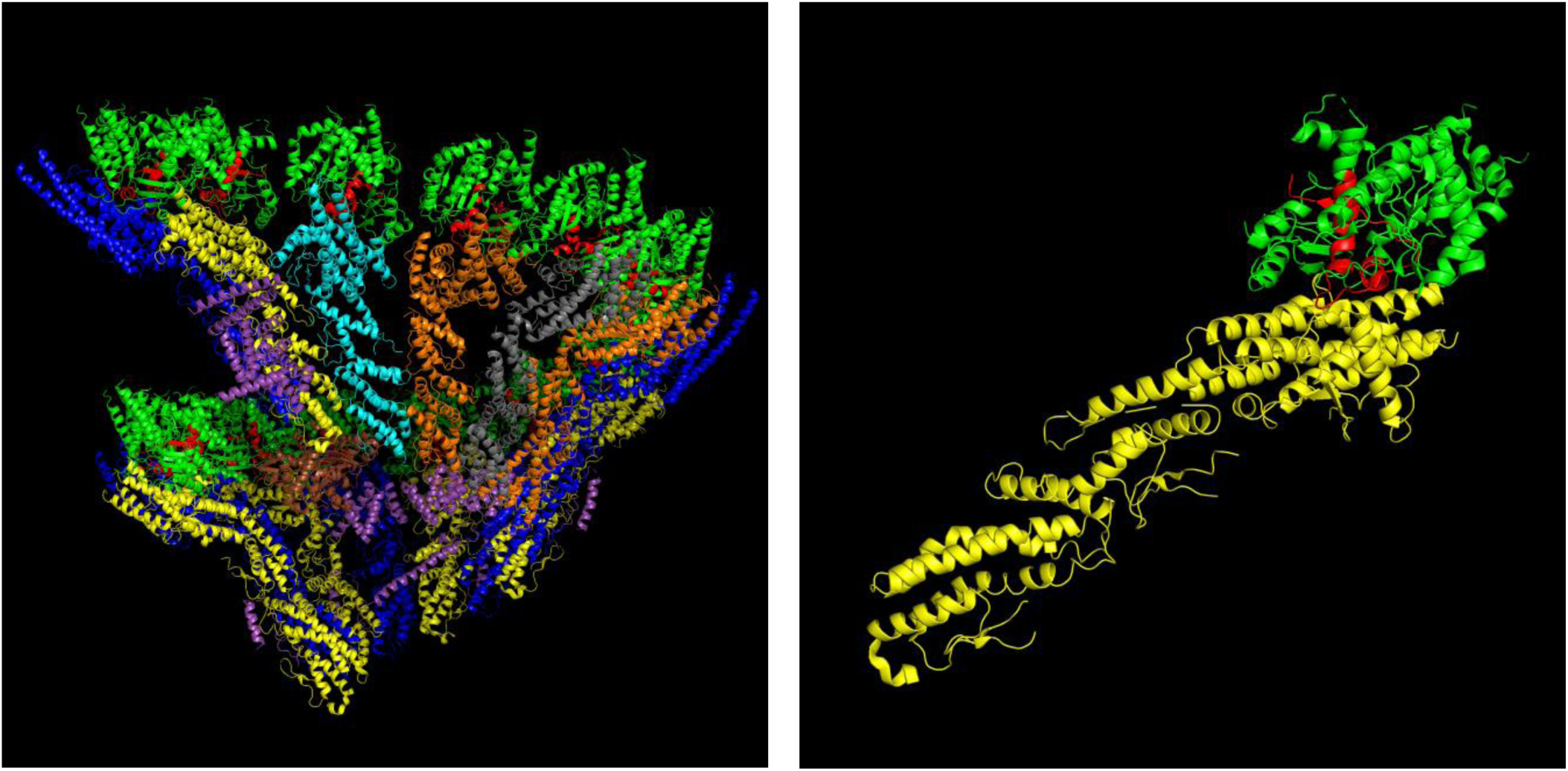
A. Visualized location of the deletion residue in a γ-TuRC. Green: gamma-tubulin, Red: amino acids 232 - 281, Yellow: GCP2, Blue: GCP3, Orange: GCP4, Grey: GCP5, Cyan: GCP6, Purple: Unassigned structures, Brown: Beta actin, B.Visualized location of the residue in a gamma-tubulin and GCP2 dimer. The residue is located on a protein-protein interface where gamma tubulin interacts with GCPs.

## Discussion

Tubulinopathy is a group of disorders characterized by a neurodevelopmental disorder and complex brain malformation including cortical dysplasia, and pachygyria. It is associated with Complex cortical dysplasia which is caused by defects of genes related to microtubule and neuronal migration ^2,20^. Tubulins compose essential parts of the microtubule and MTOC. To this day, 6 tubulin genes are reported as causal genes of tubulinopathy ^1^.

TUBG1 is one of the known tubulinopathy-related genes, which encodes tubulin gamma-1 protein. Tubulin gamma-1 protein is a component of γ-TuRC and mediates nucleation and formation of microtubules. Only autosomal dominant disease was reported related to TUBG1. Heterozygous missense variants of TUBG1 were reported by Poirier et al in 2013 ^4^. As a loss-of-function variant was not discovered from the group, authors suggested a dominant-negative effect, rather than haploinsufficiency, to be a disease-causing mechanism of variants. The suggestion can be supported by this report.

In the two probands described in this study, the clinical presentation included microcephaly, delayed psychomotor development, intellectual disability, and strabismus. Proband 1 also exhibited epilepsy. Moreover, brain MRI scanning showed the presence of subcortical band heterotopia.

All of these clinical and radiological findings align with the clinical spectrum of tubulinopathies, suggesting that the probands’ conditions may be linked to tubulinopathy.

Through exome sequencing and Sanger sequencing of the family, we have discovered a homozygous intronic variant in the probands and that the variant was inherited from consanguineous, unaffected parents in the heterozygous state. To further explore the consequences of this variant, we performed Sanger sequencing using the cDNA, which confirmed that the variant causes aberrant splicing, leading to a skipping of the exon 8, and the deletion of 150 bases in the transcript. This deletion is anticipated to cause the in-frame deletion of 50 amino acids. As the variant is resulting in an in-frame 50 amino acids deletion, it is likely to cause partial functional defects, rather than complete loss-of-function of the protein. In addition, the deleted residue is located at the external part of the protein, thus less likely to lead to misfolding or partial unfolding. Rather the deletion is likely to impact the protein-protein interaction of TUBG1 and GCPs which interaction the residue has a direct effect on. Further study is required to examine its impact.

This supports the hypothesis that the identified *TUBG1* variant could be pathogenic. However, further functional studies are needed to precisely determine the extent of this disruption and its effect on microtubule formation and neuronal migration.

The genotype-phenotype correlation has been tried to be developed by many papers, but no clear correlation has been observed^5,12^.

This phenotypic variability underscores the complex genotype-phenotype correlations in TUBG1-related disorders^4,12^. Our case report does not support the presence of related severity with gene variation type, as we could see in symptoms a severity variability has been observed between siblings, with the absence of epilepsy for the sister, with her brother was more severely ill, with more expressed neurodevelopmental delay, beside his testosterone hormone low levels, which explain the micropenis .this low level of the hormone would implicate abnormalities in the migration of basal ganglia neuron and alter their hormonal secretion levels and hypothalamic-hypophysis connection, as no abnormality of hypophysis has been observed in brain MRI expending by that the clinical presentation and expression of the disease. This clinical variability between both siblings would support the presence of other factors to be explored. However, the number of variants described remains relatively small to conclude genotype-phenotype correlation. On the other hand, the transmission mode and clinical presentation and transmission model of the disease in the presented family support the idea of the dominant negative effect suggested to be the mechanism of disease physiopathology.

The inheritance pattern observed in this case is also novel. While most *TUBG1* mutations are de novo and exhibit autosomal dominant inheritance, the homozygous nature of the variant in the probands, combined with the unaffected heterozygous parents, suggests a recessive inheritance pattern. Both parents are carriers of the variant but exhibit no clinical symptoms, indicating that one functional copy of *TUBG1* is sufficient to support normal neuronal development. This expands the understanding of *TUBG1*-related disorders and introduces a new dimension to the genetic mechanisms underlying tubulinopathies.

## Conclusion

In conclusion, we report a novel homozygous splice variant in *TUBG1* in two siblings presenting with clinical features consistent with tubulinopathy. This study is significant in expanding the known mutational spectrum of *TUBG1* and proposing a novel recessive inheritance pattern for *TUBG1*-related disorders. However, further studies are needed to confirm the pathogenicity of this variant and to clarify the genotype-phenotype relationship in tubulinopathies. Understanding the molecular mechanisms underlying *TUBG1*-related disorders will pave the way for potential therapeutic interventions, including gene therapy and personalized medicine approaches.

## Data Availability

The data that support the findings of this study are available from the corresponding author upon reasonable request.

## Declarations

### Ethics approval and consent to participate

The family consented for a genetic study approved by the relevant ethics committees and institutional review boards of the Fez University Hospital EthicsCommittee, with ethics committee approval given to this study under number : 02/2023 in accordance with the Declaration of Helsinki.

### Author contributions

Badreddine ELMAKHZEN, Yongjun SONG: Wrote the manuscript, Data analysis, figures, original manuscript writing and editing. Clinical data collection, manuscript drafting.

Yongjun SONG, Omar ASKANDER^2^ : Data analysis, WES analysis, and interpretation.

KETTANI OUSSAMA, Yasser ALI EL ASRI : Conceptualization and resources, Clinical data collection.

Yongjun SONG, Omar ASKANDER, Laila BOUGUENOUCH, ZINEB RCHIAD : Manuscript review and editing, conceptualization, and supervision.

All authors provided valuable feedback on earlier versions of the manuscript. The final manuscript was reviewed and approved by all authors.

## Acknowledgments

The authors express their gratitude to all study participants, their families, collaborating physicians,, and laboratory staff who helped with patient diagnosis, blood collection,DNA preparation, and sequencing.

